# Distinct molecular profiles characterize the spontaneous growth rate of IDHmt low-grade astrocytoma and oligodendroglioma, WHO grade 2

**DOI:** 10.1101/2025.10.21.25338454

**Authors:** Amélie Darlix, Pierre Bady, Jérémy Deverdun, Emmanuelle Le Bars, Arthur Coget, Justine Meriadec, Mathilde Carrière, Hugues Duffau, Monika E. Hegi

**Author notes:** **Corresponding Author:** Monika E. Hegi, PhD, Lausanne University Hospital, Centre Hospitalier Universitaire Vaudois, Chemin des Boveresses 155, CLE-C 306, 1066 Epalinges, Switzerland. These two authors contributed equally (Amélie Darlix, Pierre Bady).

## Abstract

**Background:** The life expectancy of patients with diffuse IDH-mutant low-grade gliomas (IDHmt LGG) WHO grade 2 ranges from 5 to over 20 years. Tumor behavior, including spontaneous growth rate, varies even within homogeneously classified subtypes of oligodendroglioma and astrocytoma. Risk-adjusted treatment strategies are needed to avoid therapy-related toxicities, without compromising outcome. The spontaneous tumor volume growth rate (TVGR) serves as a prognostic marker and predicts response to therapy. Accurate prediction of TVGR through biomarkers would enable an improved evidence-based risk management.

**Patients & Methods:** A cohort of 77 patients treated in Montpellier, France, for IDHmt LGG grade 2 (29 oligodendrogliomas, 48 astrocytomas) was analyzed (age >18 years; MRI scans and frozen tumor tissue available). DNA methylome profiling (Illumina, EPIC array) and RNA sequencing were established. TVGR was determined based on serial MRI collected over the “watch & wait” period from diagnosis to first treatment beyond surgery. Transcriptomic and methylome data were analyzed for signatures associated with TVGR using rank-rank regression followed by preranked gene set enrichment analysis.

**Results:** The median TVGR was lower in IDHmt codeleted compared to non-codeleted LGG, (0.241 year^-1^ range 0.082-0.366 vs 0.424 year^-1^ range 0.264-0.609, p<0.001). In codeleted IDHmt LGG, TVGR was associated with deregulated gene signatures for signal transduction, neuronal systems, growth factor stimulation, and neural progenitor- and stem cells. In contrast, TVGR in non-codeleted IDHmt LGG was associated predominantly with proliferation-related signatures.

**Conclusion:** Spontaneous TVGR of codeleted and non-codeleted IDHmt LGG involve distinct biological processes, suggesting possible differences in response to therapies.

**Key Points:**

- IDHmt glioma WHO grade 2 growth rate impacts outcome and response to therapy
- Molecular drivers of spontaneous growth rates are distinct between astrocytomas and oligodendrogliomas
- Possible impact on treatment response and outcome

**Importance of Study:** The life expectancy of patients with diffuse IDH-mutant low-grade gliomas (IDHmt LGG) WHO grade 2 ranges from 5 to over 20 years, with tumor behavior and growth rate varying even within homogeneous histo-molecular subtypes. Risk-adapted treatment strategies are needed to avoid therapy-related toxicities, without compromising outcome. The spontaneous tumor volume growth rate (TVGR) is a prognostic marker and predicts therapy response. We analyzed a cohort of 77 patients with IDHmt LGG, with available MRI and tumor tissue. DNA methylome profiling and RNA sequencing were performed. TVGR was calculated from serial MRI during the “watch & wait” period before any treatment beyond surgery. TVGR in non-codeleted LGG patients was mainly associated with gene signatures linked with proliferation, while it was associated with deregulated gene signatures for signal transduction, neuronal systems, growth factor stimulation, and neural progenitors and stem cells in codeleted tumors. These insights suggest possible differences in response to therapies.

## INTRODUCTION

Diffuse low-grade gliomas (LGG) WHO grade 2 are rare tumors with a yearly incidence of 1-2/10^5^ that typically affect young patients before the age of forty. These tumors are characterized by a mutation in the isocitrate dehydrogenase *(IDH)* gene 1 or 2 (IDHmt) that is associated with a CpG island methylator phenotype (CIMP) that affects expression of large sets of genes. Although slowly growing, these tumors eventually progress to higher grade, with a median overall survival (OS) of 5 to over 20 years (Ng et al., 2024). The prognosis depends on the genetic subtype in particular the presence of a codeletion of the chromosomal arms 1p and 19q (further referred to as codel) as well as clinical and radiological parameters such as tumor volume, and spontaneous growth rate. Maximal safe surgical resection is the first line treatment whenever possible, and the extent of resection has a positive prognostic impact (Ng *et al*., 2024). However, the timing and choice of further treatments including systemic treatments (cytotoxic, targeted, or immunotherapies), radiation therapy (RT) or combinations thereof are debated. Depending on prognostic factors, a “watch & wait” strategy can be proposed following tumor resection in patients with a small residual volume. When clinical and/or radiologic criteria suggest progression, additional treatments (new surgery, systemic treatment and/or RT) are initiated. The time interval between surgery / histological diagnosis and the initiation of the first medical treatment, beyond surgical resection, varies on a case-by-case basis, up to several years in selected cases. The growth rate of the tumor is a critical parameter to define the therapeutic treatment. It ranges from 1 to over 40 mm/year (median 4 mm/year), and has been shown to be an independent prognostic factor (Pallud et al., 2013), including in subgroups of molecularly-defined tumors according to the 2016 WHO classification (Leclerc et al., 2024; Roux et al., 2020). Lower growth rates have been observed in codel than in non-codel LGG (Huang et al., 2020). The growth rate also has an impact on response to treatments (Darlix et al., in preparation). To date however, the mechanisms of the underlying biology are unknown due to lack of associated molecular data.

We aimed at identifying molecular features associated the tumor volume growth rate (TVGR) in a patient cohort constituted in a single institution (Montpellier, France) where the “watch & wait” strategy is usually proposed following tumor resection. The TVGR was established based on MRI data collected over the “watch and wait” period up to the time of initiation of the first oncologic treatment with the alkylating agent temozolomide (TMZ). To gain insights into the molecular underpinnings of the natural evolution of IDHmt LGG grade 2, the tumor methylome (EPIC 850k methylation array) and transcriptome (RNA sequencing) were determined and analyzed and associations with the TVGR were evaluated.

## MEHODS

### Patient Selection

A cohort of patients (≥18 years-old) treated between 2002 and 2022 for a diffuse IDHmt low-grade (WHO grade 2) glioma was constituted at the Institut régional du Cancer Montpellier (ICM) (BDD-NO database), France, as previously described (Darlix et al., 2025). In brief, selection criteria for patient inclusion comprised, TMZ as first-line oncologic treatment after surgery/ies, availability of frozen tumor tissue (biobanque, BB-0033-00059, CRB-ICM) and brain MRIs in *Digital Imaging and Communications in Medicine* (DICOM) format collected during the “watch & wait” period (MRI performed every 3 to 6 months) from first tumor diagnosis to the initiation of TMZ treatment, defined as first intervention free survival, FIFS. Treatment initiation was based on clinical (location, clinical symptoms) and radiological criteria (large post-surgical FLAIR tumor residue or tumor FLAIR volume progression. The standard treatment schedule was as follows; TMZ, day 1 to 5, every 28 days, 150mg/m2/day for the first cycle and 200mg/m2/day for the subsequent cycles as reported (Darlix *et al*., 2025). Tumor DNA methylome data (Illumina EPIC arrays) were available for 101 patients.

Patients’ data were collected until September 19^th^, 2025. All patients provided consent for translational research. The study was approved by the local Institutional Review Board (authorization #ICM-CORT-2019-2) and the ethics committee of the canton de Vaud (protocol CER-VD 2020-02519) Lausanne, Switzerland.

### Automated determination of Tumor volumes

MRI scans in DICOM format were collected. T1-weighted (T1w) and T2-FLAIR series were converted to NIfTI format and structured according to the Brain Imaging Data Structure (BIDS). Tumors were segmented using an automated procedure (Verdier et al., 2024), which has demonstrated good performance, while significantly reducing the medical effort needed for corrections. All segmentations were then visually reviewed by three experienced neuroradiologists (AC, JM, MC) and manually corrected if necessary, using the MRIcron software. T2-FLAIR imaging and both anterior and posterior timepoints were used to maintain consistency across exams. Finally, each segmentation was rechecked by one of the three neuroradiologists who performed the corrections, ensuring that the reviewer was different from the one who initially checked the segmentation. The tumor volumes served as input to calculate the TVGR. The mean tumor diameter (MTD) was estimated from the tumor volume (mm3) using the following formula: MTD (mm) = (2 × V)^(1/3).

### Modelling tumor growth and estimation of the growth rate

The growth curves were fitted by a mixed linear model from the linearization of the exponential model for tumor volume and linear model for MTD. The model includes random intercept to take in account the lead-time bias (tumor sizes at the time of diagnosis) as proposed in Mandonnet et al. (Mandonnet et al., 2003), and random slope to estimate the tumor progression for each patient. The model was completed by the subtype (1p/19q codeletion status) and interaction effect between subtype and time as fixed effects. The variance structure of within-group errors was modeled by a power variance function in using covariates (Pinheiro and Bates., 2000).

The individual relative volume growth rate and linear diameter growth rate are given by the random and fixed slopes of the model and the coefficient related to interaction term (Heesterman et al., 2019). A bootstrap procedure was used to consolidate the estimation of the relative TVGR and the linear MDT growth rate (TDGR) by the mean of the slopes from 50 repetitions and to provide confidence intervals of TVGR and TDGR. The tumor volume doubling time is defined as ln(2)/TVGR. The mixed models were provided by the function nlme from R package nlme (Pinheiro and Bates., 2000). The bootstrap of the mixed model was obtained by the function boot_lme from R package nlraa (Archontoulis and Miguez, 2015).

### DNA methylome analyses

Establishment of the methylome data (Ilumina EPIC array) and respective preprocessing for the Montpellier cohort has been described previously (Darlix *et al*., 2025). The dataset is available under the GEO accession number GSE279950 (http://www.ncbi.nlm.nih.gov/geo/).

#### IDH mutation classification

Only IDH mutant tumors, confirmed using a methylome-based procedure (Darlix *et al*., 2025; Yang et al., 2022) were retained for this study.

#### DNA copy number assessment and 1p/19q codeletion status

DNA copy number assessments and determination of the 1p/19q codeletion status were performed using the DNA methylation data as described (Bady et al., 2018; Darlix *et al*., 2025).

#### *MGMT* promoter methylation

The DNA methylation status of the *MGMT* promoter *(MGMTp)* and the *MGMT* score (logit-transformed probability) were determined based on the EPIC data using the methylation probes, *cg12434587* and *cg12981137* and the logistic regression model MGMT-STP27 as previously described (Bady et al., 2016; Bady et al., 2012). The Confidence Interval and MGMT classifications can directly be obtained by the function *MGMTpredict* from the R package mgmtstp27 for Inifinium platforms (27k, 450k, EPIC and EPICv2, version 0.8, https://github.com/badozor/mgmtstp27).

#### Data preparation and analysis of RNA sequencing

RNA was extracted from frozen tumor tissue using the AllPrep DNA/RNA Micro Kit (Qiagen, Cat. No./ ID: 80284) as previously described (Darlix *et al*., 2025). RNA quality was assessed on a Fragment Analyzer (Agilent Technologies). The RNAs had RQNs between 6.3 and 9.7. The samples were randomized on a 96 well plate. RNA-seq libraries were prepared from 50 ng of total RNA with the Illumina Stranded mRNA Prep reagents (Illumina) using a unique dual indexing strategy, and following the official protocol automated on a Sciclone liquid handling robot (PerkinElmer). Libraries were quantified by a fluorometric method (QubIT, Life Technologies) and their quality assessed on a Fragment Analyzer (Agilent Technologies). Sequencing was performed on an Illumina NovaSeq 6000 S2 v1.5 flow cell for 100 cycles single read. Sequencing data were demultiplexed using the bcl2fastq2 Conversion Software (version 2.20, Illumina). RNA sequencing was performed at the Lausanne Genomic Technologies Facility, Center for Integrative Genomics, University of Lausanne.

The preprocessing of RNAseq data was performed following the standard pipeline and recommendations from bcbio-nextgen (version 1.0.4, http://bcbio-nextgen.readthedocs.org/en/latest/). After trimming of adapter and polyA, and standard quality control, the alignments to the human reference genome (assembly GRCh37/hg19) were performed by hisat2 aligner (version 2.1.0). The gene expression data were summarized by trimmed mean of M-values (TMM) normalized counts (R package edgeR) (Robinson et al., 2010; Robinson and Oshlack, 2010), including log-transformation and using read counts and full library size. The Variant calling analysis was performed with the software VarDict (Lai et al., 2016) and the genomic variant annotations were obtained by SNPeff (Cingolani et al., 2012). SNV not passing the quality filter and silent and synonymous mutations were excluded. The genes listed in the COSMIC database as mutated in low grade glioma (LGG) and GBM (Tate et al., 2019) were used for our analysis. In addition, the identified SNV were compared with the mutation database from TCGA for LGG and GBM (R package RTCGA.mutations). For personal privacy reasons the RNA-seq raw data will be made available upon request.

### External Datasets

External datasets comprised the grade 2 LGG dataset from The Cancer Genome Atlas characterized by IDH mutation predicted by the model from Yang et al (Yang *et al*., 2022) after quality control and filtering, (dbGaP accession number phs000178.v9.p8; http://cancergenome.nih.gov) (Brat et al., 2015)). The TCGA dataset is characterized by 94 1p-19q non-codel and 80 codel patients.

#### Pathway Analyses

Association between markers (gene and CpG-probes) and TVGR (and TDGR) was estimated by rankrank regression (Chetverikov and Wilhelm, 2024). The R package csranks provides statistical tools for estimation and inference involving ranks (the position in a ranking), and the function lmranks was used to perform rank-rank regression. The z-values from the regression provided the ranked gene lists usable for gene set enrichment analysis (GSEA).

Pathway analysis was performed by preranked gene set enrichment analysis (GSEA) based Monte-Carlo procedure (GSEA, R packages msigdbr and ClusterProfiler) using the Molecular Signatures Database (MSigDB, v7.5.1, updated 2022 collections of H hallmark genes sets and C2 curated gene sets, (Subramanian et al., 2005). The analyses were provided using function from R package clusterprofiler (Xu et al., 2024). Gene-sets with Bonferroni adjusted P-values⍰≤⍰0.05 were considered significant. The averaged expression and DNA methylation for selected pathway were weighted by their total inertia for the simultaneous heatmap representation as used in MFA (multiple factor analysis (Escofier and Pagès, 1994) *the best k is selected according to the Calinski-Harabasz criterion from R package vegan* (Chessel et al., 2004). The annotations of the selected pathways were completed by a text mining procedure.

The selected pathways were described by additional annotation from Reactome (if available) and textual analysis (e.g. pathway including the term “repair”). Pearson and Spearman correlation give information on the strongness and direction of the association between TVGR and the pathway (averaged expression or averaged DNA methylation).

The Importance of the pathways in the prediction of the TVGR was individually estimated for each pathway by r-squared coefficient from partial least square regression (PLS) of the TVGR on the gene expression (or DNA methylation probe).

### Statistical analyses

For the continuous variables, Wilcoxon test (t) or Kruskall and Wallis test (a) were used to test the differences between two or more groups. The independence between qualitative variables and groups was tested with Pearson’s Chi-squared with Yates’ continuity correction or based on permutation test.

The confidence intervals for proportion are given by exact binomial procedure (confidence level = 0.95). Survival univariate and multivariate models were computed by Cox proportional hazards regression model (Therneau and Grambsch, 2000). Analyses and graphical representations were performed using R-4.4.3 and the R package rms and survival (R2014, rms2014 and survival 2014). Similarity between matrices was evaluated by RV (vectorial correlation) coefficient (values between 0 and 1) (Escoufier, 1973), using R package ade4 for pairwise RV coefficient permutation tests (Chessel *et al*., 2004) and computation of the MFA (Chessel *et al*., 2004).

## RESULTS

### Patient cohort

#### Tumor growth rate is higher in non-codel than in codel IDHmt LGG WHO grade 2

A total of 88 patients (codel, n=30; non-codel, n=58) corresponding to 441 MRI scans in DICOM format over the “wait & watch” period up to the time of treatment initiation with TMZ (defined as First Intervention-Free Survival, FIFS) was initially selected (see sample flow in Suppl Figure S1A). Among them, 78 also had matching molecular data. After excluding 1 patient with low QC data, 77 patients were included in the analyses (codel, n=29; non-codel, n=48). Their baseline characteristics are summarized in Table 1.

**Table 1.** Baseline description by LGG subtype, codel and non-codel.

| Variable | N | codel<br>N = 29<sup>1</sup> | non-codel<br>N = 48<sup>1</sup> | p-value<sup>2</sup> |
| --- | --- | --- | --- | --- |
| **Median age at diagnosis** | 77 | 40 (34, 50) | 33 (29, 39) | 0.003 |
| **Biological sex** | 77 |  |  | 0.023 |
| F |  | 16 (55%) | 14 (29%) |  |
| M |  | 13 (45%) | 34 (71%) |  |
| **Tumor location** |  |  |  |  |
| Frontal | 77 | 21 (72%) | 31 (65%) | 0.5 |
| Occipital | 77 | 1 (3.4%) | 3 (6.3) | >0.9 |
| Temporal | 77 | 9 (31%) | 21 (44%) | 0.3 |
| Parietal | 77 | 6 (21%) | 8 (17%) | 0.7 |
| Insular | 77 | 7 (24%) | 18 (38%) | 0.2 |
| Basal Ganglia | 77 | 0 | 1 (2.1%) | >0.9 |
| Corpus Callosum | 77 | 2 (6.9%) | 1 (2.1%) | 0.6 |
| **Surgery type** | 77 |  |  | 0.6 |
| Biopsy |  | 2 (6.9%) | 2 (4.2%) |  |
| Surgical resection |  | 27 (93%) | 46 (96%) |  |
| **Performannce status** | 77 |  |  | 0.8 |
| 0 |  | 12 (41%) | 19 (40%) |  |
| 1 |  | 17 (59%) | 27 (56%) |  |
| 2 |  | 0 (0%) | 2 (4.2%) |  |
| **Re-resection** | 77 |  |  | 0.5 |
| no re-resection |  | 21 (72%) | 31 (65%) |  |
| re-resection |  | 8 (28%) | 17 (35%) |  |
| **MRIs/patient** | 77 | 6 (3, 8) | 4 (3, 5) | 0.005 |
| **Time from diagnosis to** |  |  |  |  |
| **<sup>3</sup>TMZ-trt, months** | 77 | 53 (29, 70) | 33 (20, 63) | 0.075 |
| **MGMT-STP27 score** | 77 | 3.61 (3.13, 4.68) | 2.24 (0.69, 2.82) | <0.001 |

<sup>1</sup> Median (Q1, Q3); n (%)
<sup>2</sup> Wilcoxon rank sum test; Pearson's Chi-squared test; Wilcoxon rank sum exact test
<sup>3</sup> *Abbreviations:* N, number; TMZ-trt, TMZ treatment; MRI, Magnetic resonance imaging

The determined tumor volumes served as input to calculate the tumor growth rate.

The median TVGR was lower in IDHmt codel LGG (oligodendroglioma) than for the non-codel (astrocytoma) as expected, 0.241 year^-1^ (range, 0.082 to 0.366) versus 0.424 year^-1^ (range, 0.264 to 0.609) (Figure 1, Table 2). The corresponding values for the TDGR are available in Table 2 and illustrated in Suppl Figure S2. The correlation coefficient between the estimation of TVGR and TDGR was 0.95 (Spearman). The median tumor volume doubling time was 2.876 years (range 1.892 to 8.425) for codel and 1.636 years (range, 1.138 to 2.629) for non-codel. The quality of the model and the estimated TVGR are visualized in Suppl Figure S3. Estimations of TVGR by patient are summarized in Suppl Table S1, and respective information for the TDGR in Suppl Table S2.

**Table 2.** Tumor Growth Rate stratified by LGG subtype, codel and non-codel.

| Variable | N | codel<br>N = 29<sup>1</sup> | non-codel<br>N = 48<sup>1</sup> | p-value<sup>2</sup> |
| --- | --- | --- | --- | --- |
| **TVGR, year<sup>-1</sup>** | 77 | 0.24 (0.19, 0.26) | 0.42 (0.40, 0.46) | <0.001 |
| **TDGR, mm/year** | 77 | 2.88 (1.97, 3.41) | 5.09 (4.57, 5.76) | <0.001 |
| **TVDT, years** | 77 | 2.88 (2.65, 3.58) | 1.64 (1.49, 1.75) | <0.001 |

<sup>1</sup> Median (Q1, Q3)<sup>2</sup> Wilcoxon rank sum exact test
*Abbreviations:* N, number; TVGR, tumor volume growth rate; TVDT, TDGR, tumor diameter growth rate; TVDT, Tumor volume doubling time.

**Figure 1.**
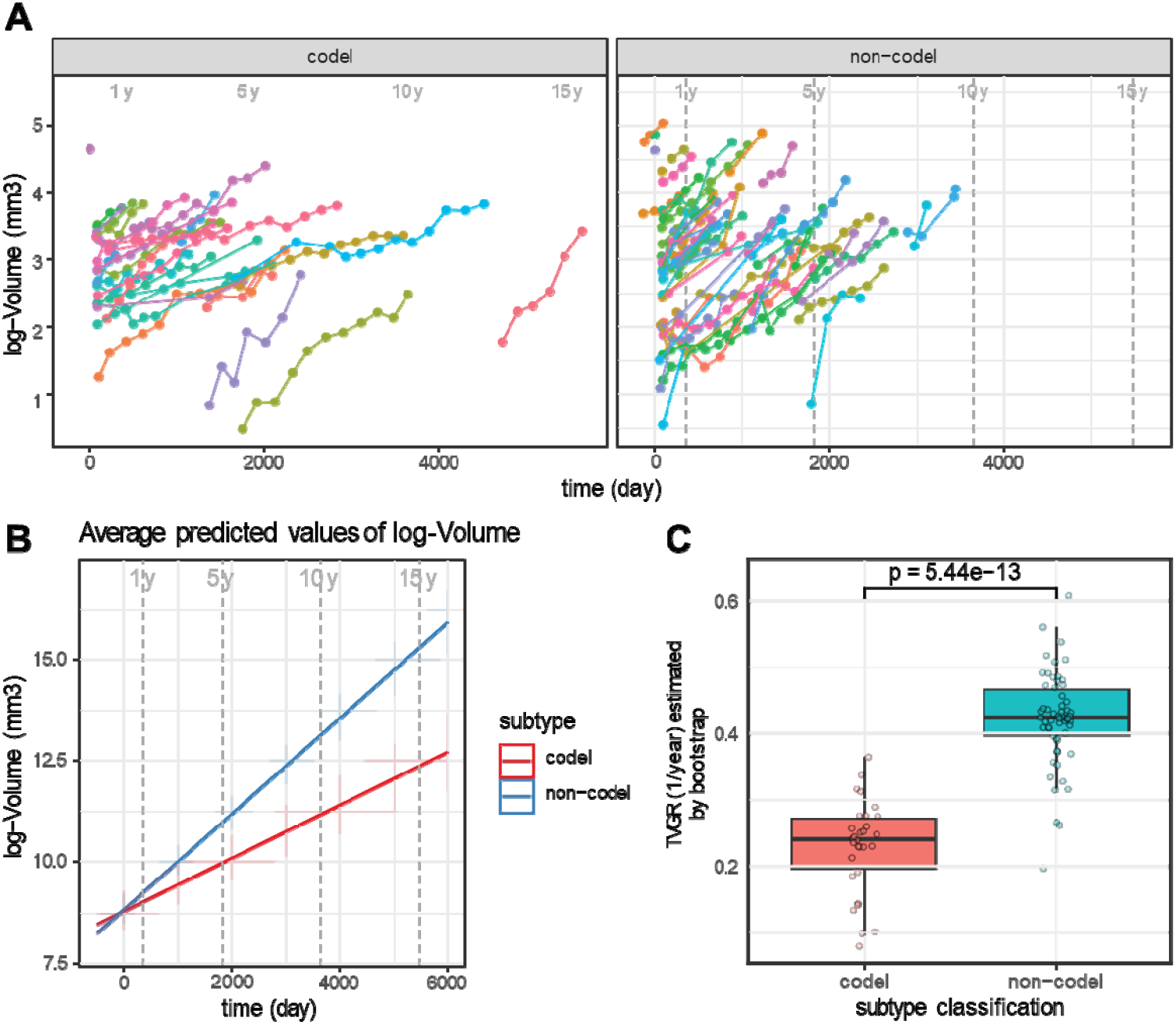
Tumor Volume Growth Rate. **A** Representation of the tumor volume (mm3) in function of the time (t=0, time of diagnosis) by subtype (codel, n=30; non-codel, n=58). B Average predicted log volume by subtype, the shaded areas correspond to the 95% confidence interval. **C** Mean Tumor Volume Growth Rate (TVGR) estimated by bootstrap in codeleted and non-codeleted LGG, visualized in a Box plot as mean TVGR/year by subtype. The corresponding analyses with the Tumor Diameter Growth Rate (TDGR) are illustrated in Supplementary Figure S2.

#### Distinct sets of molecular pathways are associated with increased TVGR in non-codel versus codel IDHmt LGG

For 77 of these patients with IDHmt LGG matching molecular data was available comprising the DNA methylome (Ilumina EPIC, 850k) and the transcriptome RNAseq. The association between markers (gene transcripts; CpG-probes; copy number variation, CNV) and the TVGR by rank-rank regression yielded gene lists that served as input into the preranked gene set enrichment analyses (GSEA) using the Molecular Signatures Database (Subramanian *et al*., 2005) (See flow of analyses, Suppl Figure S1B).

When analyzing codel (n=29) and non-codel (n=48) IDHmt LGG together for molecular associations with the TVGR, the main features of the extracted significant pathways characterized the subtype as determined by variation partitioning. A part was explained by the interaction of the subtype and the TVGR, while the variation explaining the growth rate was very low. This was true using either functional DNA methylation from promoter regions, gene expression (RNA-seq) or CNV (DNA methylation data derived) as illustrated in a heatmap (Suppl Figure S4). This was not a surprising result, emphasizing that codel and non-codel IDHmt LGG are molecularly and biologically distinct diseases.

Subsequently the analyses were performed by subtype, non-codel and codel, separately, using the respective preranked gene lists as input. The resulting significant GSEA-derived pathways associated with the TVGR were summarized in heatmaps representing pathway activities, defined as mean expression levels of the respective genes by tumor sample (Figure 2A, B). The corresponding pathways are listed in Suppl Tables S3 and S4 for non-codel and codel samples, respectively (gene expression, functional DNA methylation, and CNV). For non-codel IDHmt LGG, 219 gene signatures were found significantly associated with the TVGR. They were dominated by proliferation related signatures (cell cycle, DNA repair, DNA replication, and transcription) (Figure 2A, Suppl Table S3). Pathways negatively correlated with the TVGR comprised neural subtype related, and markers for cell identity and differentiation, e.g. neuronal differentiation. For the codel IDHmt LGG, 54 pathways were associated with the TVGR. The main features of the selected pathways comprised signal transduction, e.g. across synapses, neuronal systems, growth factor stimulation, and epigenetic features at genes related to development, differentiation, and cell identity of neural progenitor cells and stem cells (Mikkelsen et al., 2007). The top 30 pathways of the non-codeleted and codeleted tumors are illustrated in ridge plots in Supplementary Figure S5. A small set of 11 pathways overlapped in the analyses of the two subtypes, comprising transcription factor and growth factor related pathways. However, for all 11 pathways the association with the respective TVGR was in the opposite direction. A negative correlation was associated for 9 pathways in in the non-codel, while they were positively correlated in the codel, and vice versa for the other 2 pathways, as illustrated in ridge plots for the 11 common pathways for both LGG subtypes separately (Suppl Figure S6; Table S3, S4).

**Figure 2.**
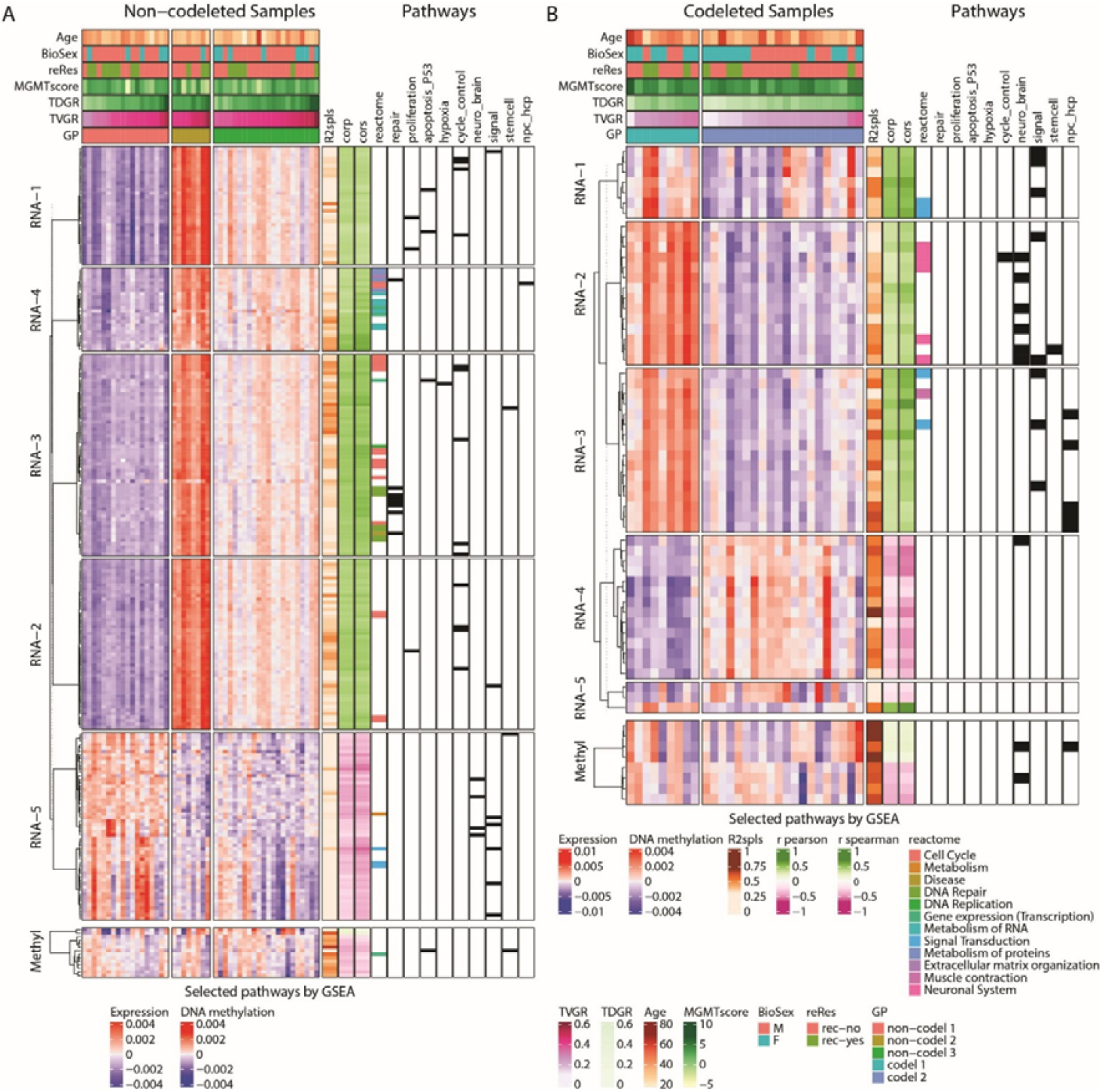
Pathways associated with TVGR for 1p/19q non-codeleted and codeleted IDHmt LGG. Pathways selected by GSEA for their associations with the estimated tumor volume growth rate. (TVGR) before the oncologic treatment intervention (TMZ), illustrated in a heatmap by LGG subtype; non-codel **(A)** and codel **(B)**. The respective gene expression and DNA methylation normalized datasets were weighted by their total inertia for the simultaneous heatmap representation, reflecting pathway activity, as used in multiple factor analysis (MFA) beforehand. The corresponding dendrograms are based on Euclidean distance and Ward’s classification for the non-codeleted **(A)** and codeleted (B) datasets. Pathway annotations comprise the associations with the TVGR (R2spls, r-squared from SPLS [sparse partial least squares]; corp, Pearson’s correlation; cors, Spearman’s correlation. Further pathway annotations are illustrated with a color code for the reactom, and in black pathways selected for recurrent signatures. Sample annotations comprise the GP, molecular group partition, TVGR, TDGR, MGMT-STP27score; reRes, reresection(s); BiolSex, biological sex, age at diagnosis.

Few pathways were associated with functional methylation in both LGG subtypes. In the non-codel IDHmt LGG, pathways were dominated by negative correlation with DNA repair pathways (3/14) and oligodendrocyte markers or differentiation associated pathways (3/14). In the codel IDHmt LGG the functional methylation-selected pathways (n=8) comprised development, differentiation in the brain.

Few signatures emerged based on CNVs. Two signatures (of 9) in the non-codel tumors were positively correlated with the TVGR and reflected amplified regions on CHR 7Q21_22, and CHR 7P22. Both signatures were also on the short list (2 of 4) for the codel group, but with the opposite (negative) correlation (Suppl Table S3, S4).

#### Transcriptome based prediction of tumor growth rate and outcome

Next, we aimed at identifying tumor subgroups with distinct pathway activities explaining the TVGR. For non-codel IDHmt LGG, partition into three molecular subgroups were proposed and two for the codel (K-means partitioning using Calinski criterion to detect the optimal partition) (Caliński and Harabasz, 1974) (Figure 2A, B, Suppl Figure S7). In both, the non-codel and the codel IDHmt tumors, the association of the group partition (GP) with the TVGR was significantly different (Figure 3A, B). The corresponding Kaplan Meyer analysis testing the GP with outcome suggested for the non-codel a significant difference for the FIFS (p=0.018, log-rank, Figure 3C) and a significant difference for overall survival (Suppl Table S5). For the codel IDHmt LGG cohort, no difference (p=0.36, log-rank) was observed for the association of the GP and FIFS (Figure 3D) (Table S5). However, the small sample size of the subgroups precludes any strong statement (n=9, n=20). The direct association of the TVGR with FIFS was significant in the non-codel patient cohort (p=0.019) and also significant for overall survival (Table S6). In the codel patients the direct association of the TVGR with FIFS did not reach significance (p=0.130). The overall survival data is immature for the codel patients (79% alive, 23/29). Of note, the TVGR was significantly associated with PFS after TMZ treatment, defined as the time interval from TMZ treatment initiation to next treatment for non-codel and codel patients. Outcome measures in association with the TVGR and the GP are summarized in Suppl Tables S5 and S6, respectively.

**Figure 3.**
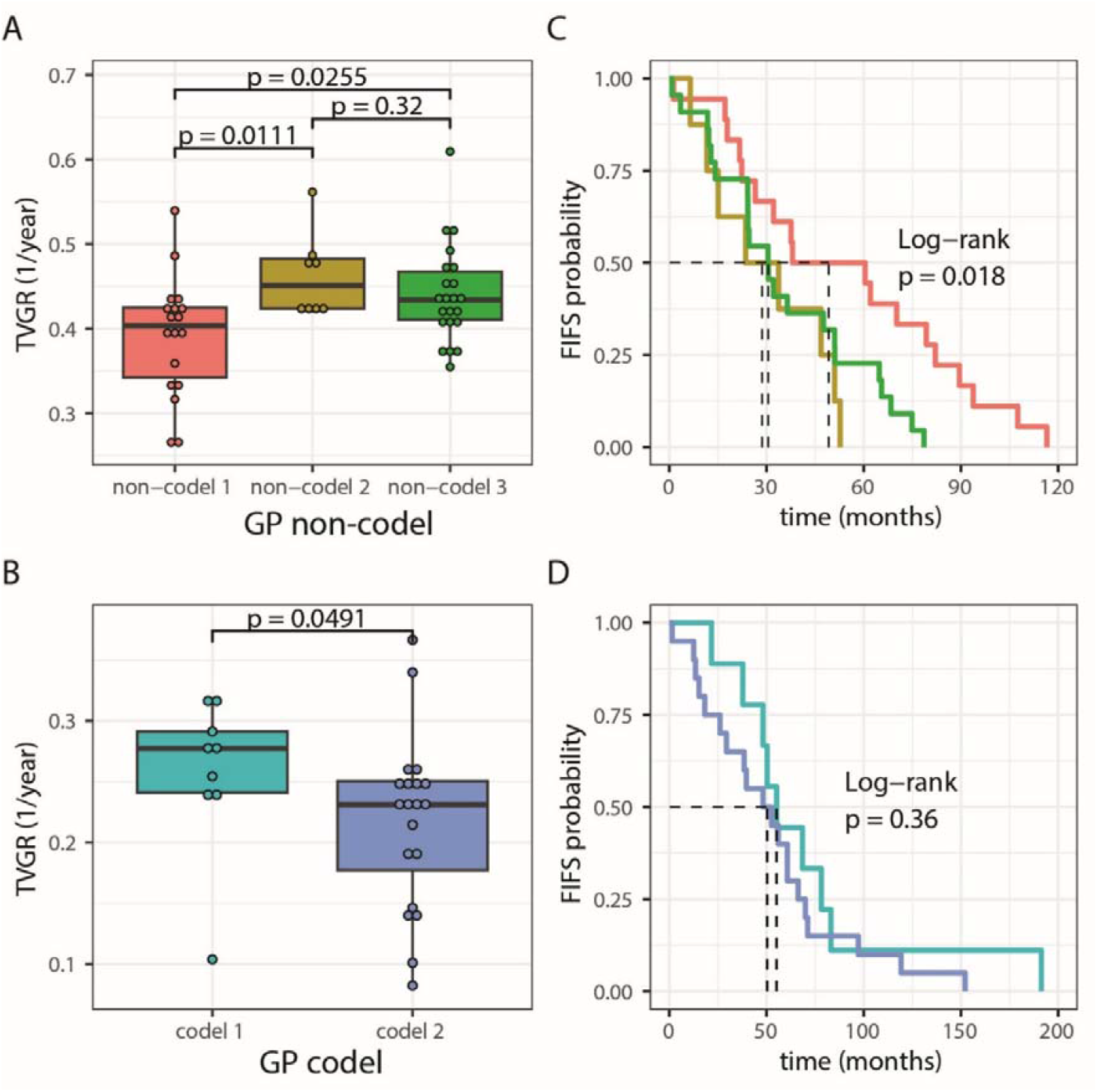
Association of molecular subgroups with first-intervention free survival (FIFS) Boxplot representation of TVGR in function of the molecular subgroups (GP, group partitions) defined by cascade K-means of pathway expression for non-codeleted (A) and codelted (B) patients. The association between first-intervention free survival (FIFS) with the GP groups are represented by Kaplan-Meier plot and tested by log-rank test for non-codeleted (C) and codeleted (D) patients.

Patients with re-resections were not enriched in any of the molecular subgroups in neither of the two glioma subtypes, non-codel or codel (Figure 2A, B, Fisher’s Exact test, p=0.5192 for non-codel and p=0.6749 for codel).

#### Evaluation of TVGR related signatures in the TCGA-LGG (IDHmt, grade 2) data set

We determined the TVGR associated expression and methylation signatures extracted in this study in the TCGA-LGG IDHmt, grade 2 samples (n=174), stratified by the codel status. Comparison to the respective data structures in the Montpellier dataset showed high similarity, as illustrated in a heatmap in Suppl Figure S7. The respective similarity measures were high, with RV coefficients of 0.91 for the expression signatures in the non-codel and 0.83 for the codel subpopulations. Similarly, the methylation-based signatures exerted high RV coefficients of 0.93 and 0.92 in non-codel and codel, respectively (Figure S7 E-H). P-values, 0.01 with 99 repetitions for all comparisons.

#### (Epi)genetic alterations

We tested the presence of (epi)genetic alterations reported to be associated with progression to higher tumor grade or malignant progression after treatment (Ghisai et al., 2024; Malta et al., 2017; Malta et al., 2024; Shirahata et al., 2018). None of the tumor samples displayed a CIMP-low phenotype and only one non-codel IDHmt LGG had a probable homozygous deletion of *CDKN2A/B*. Mutation analyses using the RNA-seq data, for mutations reported relevant in IDHmut LGG, showed a good concordance for *IDH1or 2* mutations. The next most commonly mutated gene was *TP53*, followed by mutations in *PIK3CA* and *PIK3R1*. Other mutations were rare (Suppl Figure S8). *ATRX* mutations were underreported, however, RNA based mutation analyses have limited sensitivity as they depend on transcript levels.

## Discussion

In patients with IDHmt gliomas grade 2, tumor behavior is heterogeneous within homogeneous subtypes of tumors according to the WHO 2016 classification, in terms of outcome and spontaneous growth rate (Leclerc *et al*., 2024; Roux *et al*., 2020). Growth rate is associated with survival (Pallud *et al*., 2013) and response to treatments (Darlix et al., in preparation). In this context, understanding its biological correlates is of upmost importance, to adapt the treatment strategy at the individual level. In this study, we aimed at identifying molecular features associated with the spontaneous TVGR in a cohort of patients diagnosed with IDHmt non-codel and codel LGG, to uncover, and maybe, in the near future, act upon molecular characteristics that are associated with increased growth rate. The TVGR was established based on MRI data collected over the “wait & watch” period from tumor diagnosis up to the time of treatment initiation with TMZ (FIFS). The median TVGR was lower in IDHmt codel LGG (oligodendroglioma) than the non-codel (astrocytoma) as expected (Bhatia et al., 2024; Huang *et al*., 2020). The TVGR per se had a significant association with FIFS in the IDHmt non-codel patient cohort, and a borderline significant association in the IDHmt codel cohort.

Insights into the molecular analyses revealed that gene signatures associated with the TVGR were dominated by upregulation of proliferation related pathways, such as cell cycle, DNA repair, DNA replication, and transcription in the non-codel IDHmt LGG. Respective expression profiles suggested 3 distinct molecular patient subgroups that were significantly associated with FIFS. In contrast, in codel IDHmt LGGs the association of the transcriptome with the TVGR yielded different deregulated sets of gene signatures reflecting multiple pathways, such as signal transduction (e.g. across synapses), neuronal systems, growth factor stimulation, and differentiation related signatures of neural progenitor cells and stem cells, but not proliferation related signatures. This may suggest that the TVGRs of the spontaneous evolution of non-codel and codel IDHmt LGG grade 2 are associated with and driven by distinct biological processes that may imply differences in their response to specific therapies. We could confirm the identified molecular signatures associated with the TVGR in the Montpellier cohorts in TCGA-LGG (grade2) dataset for the IDHmt non-codel and codel LGGs, respectively. In this context, it is noteworthy that an IDHm targeted drug, which reduces the production of the oncometabolite 2-hydroxy glutarate, influenced the lineage differentiation state of tumor cells in IDHmt codel LGG of patients who responded to the drug. At the time of tumor resection, these patients had been undergoing treatment for 4 weeks as part of a phase I clinical trial with the mutant IDH1/2 inhibitor ivosidenib (Spitzer et al., 2024).

There are some limitations to this study. While we were able to validate the gene signatures in the TCGA-LGG cohorts, we could not evaluate the association with growth rate and outcome. The patients in the Montpellier cohort were selected to evaluate the spontaneous TVGR during the “watch & wait” period, from diagnosis to initiation of any oncologic treatment (apart surgery), defined as First intervention-free survival (FIFS). By definition of the inclusion criteria, all patients included reached this event. This contrasts with the outcome information available for patients in the IDHmt grade 2 TCGA-LGG cohort; where the proportion of events and definition of a Progression Free Interval (PFI, time from diagnosis to the occurrence of any new tumor event) is different, 0.56 (34 of 60) for non-codel and 0.23 (15 of 65) for codel (Lin et al., 2024). Therefore, outcome data are not directly comparable and the association of the molecular profiles with the FIFS cannot be validated and warrants further studies.

Taken together, the TVGR of the spontaneous evolution of non-codel IDHmt and codel LGG is associated with distinct biological processes. This implies that they may respond differentially to therapies. Insights into the biological mechanisms associated with the tumor growth rate may facilitate risk-optimized choices, together with clinical parameters, such as tumor location and size, and patient preference, when additional treatments are required.

## Supporting information

Suppl_Tabels&Figures

## Ethics declaration

All patients provided consent for translational research. The study was approved by the local Institutional Review Board of the Institut régional du Cancer Montpellier (ICM), France (authorization #ICM-CORT-2019-2) and the ethics committee of the canton de Vaud (protocol CER-VD 2020-02519) Lausanne, Switzerland.

## Acknowledgments

Many thanks to the Biobanking Laboratory and the Molecular Biology Laboratory at the Institute of Pathology at the Lausanne University Hospital for ribonucleic acid extraction, and the Lausanne Genomic Technologies Facility, Center for Integrative Genomics, University of Lausanne for RNA sequencing. This work has benefited from the facilities and expertise of the CRB Collection (NEUROLOGIE) of the University Hospital of Montpellier, France (www.chu-montpellier.fr). The results presented here are in part based upon data generated by the TCGA Research Network: https://www.cancer.gov/tcga.

## Authorship statement

Conception and design: M.E.H, A.D. P.B., J.D. Acquisition of data (patient data, review of radiology): A.D., E.L.B., J.M., M.C., A.C., H.D.). Analysis and interpretation of data (eg, statistical analysis, biostatistics, and computational analysis): M.E.H., P.B., A.D. Manuscript writing: M.EH., P.B., A.D. Review, and/or revision of the manuscript: all coauthors. Study supervision: M.E.H., A.D.

## Data availability

Due to privacy reasons the RNA sequencing data will be made available upon reasonable request under a Data Transfer Agreement. The methylome data from the Montpellier cohort is available under the GEO accession number GSE279950

(https://www.ncbi.nlm.nih.gov/geo/query/acc.cgi?acc=GSE279950). The external datasets used, comprised methylome and RNA sequencing (Level 3) data from the LGG dataset of The Cancer Genome Atlas dbGaP accession number phs000178.v9.p8; http://cancergenome.nih.gov).

