## Supplementary material for "Distinct molecular profiles characterize the spontaneous growth rate of IDHmt low-grade astrocytoma and oligodendroglioma, WHO grade 2": Suppl_Tabels&Figures

Contents Supplement

### Supplementary Tables

#### Table S1. Tumor Volume Growth Rate (TVGR) estimated by bootstrap procedure.

#### Table S2. Tumor Diameter Growth Rate (TDGR) estimated by bootstrap procedure.

#### Table S3. Results of GSEAs for non-codeleted (CNV, Expression, DNA methylation) from marker ranking based on the rank-rank linear models.

#### Table S4. Results of GSEAs for codeleted (CNV, Expression, DNA methylation) from marker ranking based on the rank-rank linear models.

#### Table S5. Molecular Group Partition (GP) subtypes and survival analysis from cox regression models

|  | **Characteristic** | **N** | **N Events** | **log(HR)** | **95% CI** | **p-value** |
| --- | --- | --- | --- | --- | --- | --- |
|  | **Non-Codel** | | | | | |
|  | GP, FIFS | 48 | 48 |  |  |  |
|  | Non-codel-1 |  |  | - | - | - |
|  | Non-codel -2 |  |  | 3.24 | 1.27, 8.31 | 0.014 |
|  | Non-codel -3 |  |  | 2.38 | 1.16, 4.88 | 0.019 |
| GP, ^2^diagn to NxtTr after TMZ | | 48 | 46 |  |  |  |
|  | Non-codel-1 |  |  | - | - |  |
|  | Non-codel -2 |  |  | 3.49 | 1.36, 8.99 | 0.01 |
|  | Non-codel -3 |  |  | 2.14 | 0.99,4.61 | 0.052 |
|  | GP, ^3^start TMZ to NxtTr | 48 | 46 |  |  |  |
|  | Non-codel 1 |  |  | - | - |  |
|  | Non-codel -2 |  |  | 2.21 | 0.94, 5.23 | 0.07 |
|  | Non-codel -3 |  |  | 0.96 | 0.46, 1.88 | 0.9 |
|  | GP, overall survival | 48 | 39 |  |  |  |
| Non-codel 1 | |  |  | - | - |  |
| Non-codel -2 | |  |  | 1.2 | 0.29, 2.1 | 0.010 |
| Non-codel -3 | |  |  | 0.52 | -0.23, 1.3 | 0.2 |
|  | **Codel** |  |  |  |  |  |
| GP, ^1^FIFS | | 29 | 29 |  |  |  |
| Codel-1 | |  |  | - | - |  |
| Codel-2 | |  |  | 0.38 | -0.45, 1.2 | 0.4 |
| GP, ^2^diagn to NxtTr after TMZ | | 29 | 23 |  |  |  |
| Codel-1 | |  |  | - | - |  |
| Codel-2 | |  |  | 0.01 | 0.91, 0.885 | >0.9 |
| GP, ^3^start TMZ to NxtTr | | 29 | 23 |  |  |  |
| Codel-1 | |  |  | - | - |  |
| Codel-2 | |  |  | 0.33 | -0.56, 1.2 | 0.5 |
| GP, overall survival | | 29 | 6 |  |  |  |
| Codel-1 | |  |  | - | - |  |
| Codel-2 | |  |  | -0.11 | -1.7,.1.5, | 0.9 |
|  | Abbreviations: GP; Molecular Group Partition; CI = Confidence Interval, HR = Hazard Ratio; FIFS, First Intervention-Free Survival | | | | | |
|  | ^1^Time interval from diagnosis to start of TMZ, FIFS | | |  |  |  |
|  | ^2^Time interval from diagnosis to next treatment after TMZ | | |  |  |  |
|  | ^3^Time interval from start TMZ to next treatment | |  |  |  |  |

#### Table S6. Spontaneous Tumor Volume Growth Rate and Survival analysis from cox regression models

|  | **Characteristic** | **N** | **N Events** | **log(HR)** | **95% CI** | **p-value** |
| --- | --- | --- | --- | --- | --- | --- |
|  | **Non-Codel** | | | | | |
|  | TVGR, ^1^diagn to start TMZ; FIFS | 48 | 48 | 4.1 | 0. 67, 7.6 | 0.019 |
|  | TVGR, ^2^diagn to NxtTr after TMZ | 48 | 46 | 4.9 | 0.84, 9.0 | 0.018 |
|  | TVGR, ^3^start TMZ to NxtTr | 48 | 46 | 6.4 | 2.7,10 | <0.001 |
| TVGR, overall survival | | 48 | 39 | 6.1 | 1.8, 10 | 0.005 |
|  | **Codel** |  |  |  |  |  |
|  | TVGR, ^1^diagn to start TMZ; FIFS | 29 | 29 | 4.3 | -1.2, 9.7 | 0.13 |
|  | TVGR, ^2^diagn to NxtTr after TMZ | 29 | 23 | 5.4 | -0.39, 11 | 0.068 |
|  | TVGR, ^3^start TMZ to NxtTr | 29 | 23 | 6.7 | 0.55, 13 | 0.033 |
|  | TVGR, ^4^overall survival | 29 | 6 | 5.3 | -6.3, 21 | 0.3 |
|  | Abbreviations: TVGR, Tumor Volume Growth Rate; CI = Confidence Interval, HR = Hazard Ratio; FIFS, First Intervention-Free Survival | | | | | |
|  | ^1^Time interval from diagnosis to start of TMZ, FIFS | | |  |  |  |
|  | ^2^Time interval from diagnosis to next treatment after TMZ | | |  |  |  |
|  | ^3^Time interval from start TMZ to next treatment | |  |  |  |  |
|  | ^4^Time interval from diagnosis | |  |  |  |  |

### Supplementary Figures

A

B

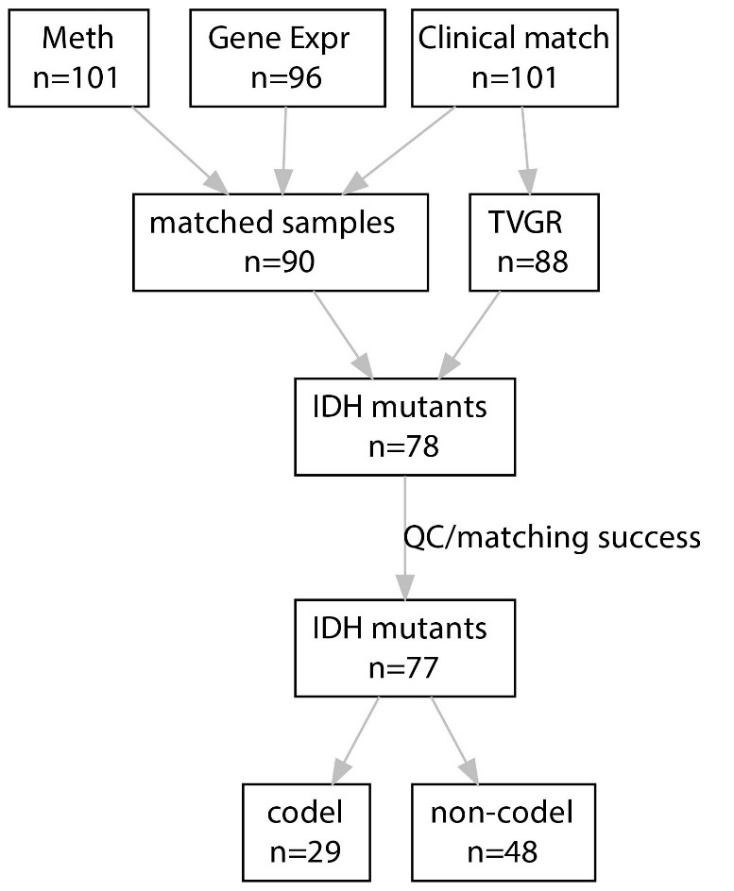

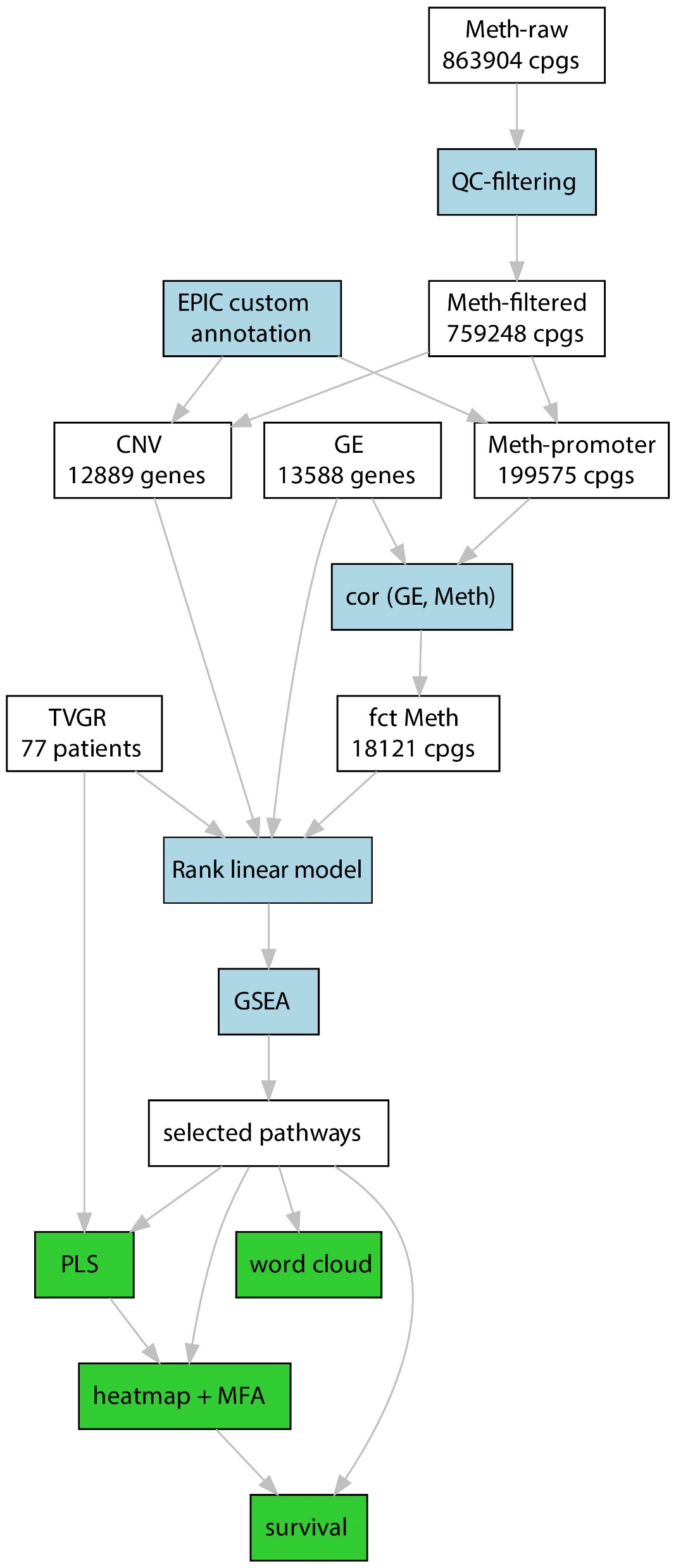

#### Figure S1. Flow diagrams for sample selection (A) and analyses (B).

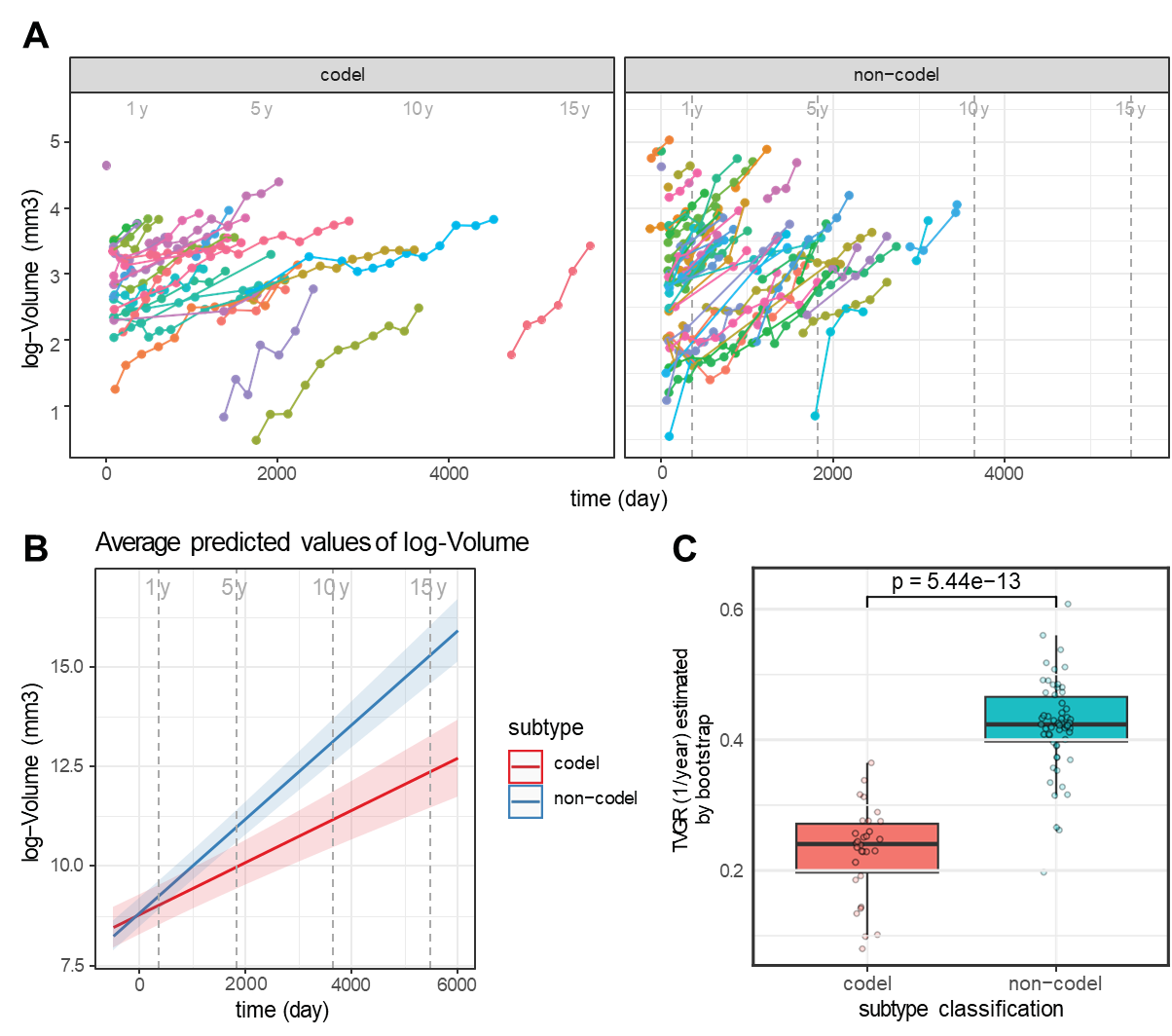

#### Figure S2. Tumor Diameter Growth Rate (TDGR)

**A** Representation of the tumor diameter (mm) in function of the time (t=0, time of diagnosis) by subtype (codel, n=30; non-codel, n=58). **B** Average predicted median diameter by subtype, the shaded areas correspond to the 95% confidence interval. **C** Mean Tumor Diameter Growth Rate (TDGR) estimated by bootstrap in codeleted and non-codeleted LGG, visualized in a Box plot as mean TDGR (mm/year) by subtype.

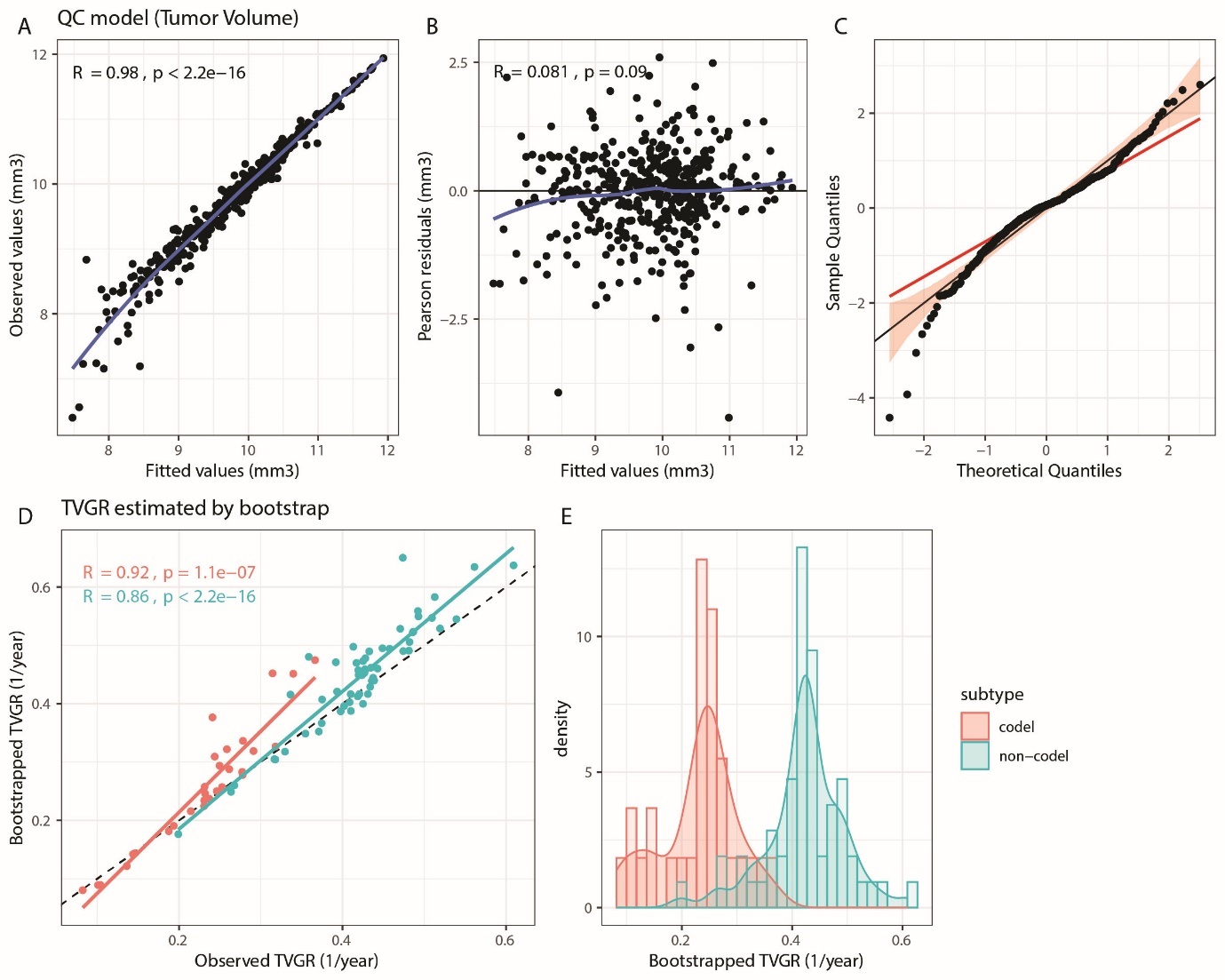

#### Figure S3. Quality control of mixed linear model and estimation for Tumor Volume Growth Rate (TVGR) by bootstrap.

Representation of the fitted against observed values for the tumor volume (A). The Pearson residuals are represented against fitted values (B) and residuals are plotted against theoretical quantiles from Normal distribution (C). The estimation of the TVGR is obtained by the mean of the slopes of bootstrapped mixed linear model from 50 repetitions. TVGR from bootstrap procedure are compared to the observed values (D) by 1p/19q codeletion subtype. The distribution TVGR are given in function of the 1p/19q codeletion subtype by density plot (E).

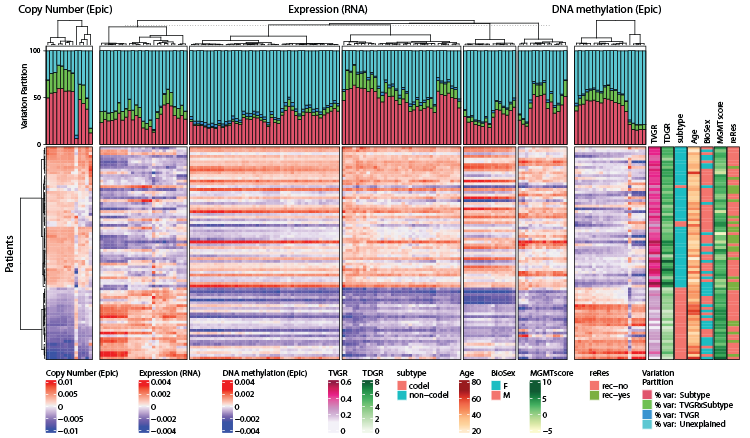

#### Figure S4. Heatmap of TVGR associated pathway activities extracted for the entire IDHmt LGG population (codel & non-codel).

The heatmap is based on the averaged gene expression, CNV and DNA methylation of the significant pathways emerging from GSEA (p ≤ 0.05). The CNV, DNA methylation and gene expression normalized datasets were weighted by their total inertia for the simultaneous heatmap representation as used in multiple factor analysis (MFA) beforehand. The dendrograms are based on Euclidean distance and Ward’s classification. The samples are clustered by copy number variation. The contribution of subtype and TVGR to explain the variance is evaluated by variation partitioning represented by bar plots of variation fractions (percentage) for the two supplementary variables (variation explained by subtype, TVGR, and interaction between subtype and TVGR). Sample annotations comprise the TVGR, TDGR, subtype by 1p/19q codeletion status, age, time from diagnosis to first oncologic treatment initiation (FIFS, first Intervention Free Survival), MGMT-STP27score, surgery type, reresections (reRes), sample ID.

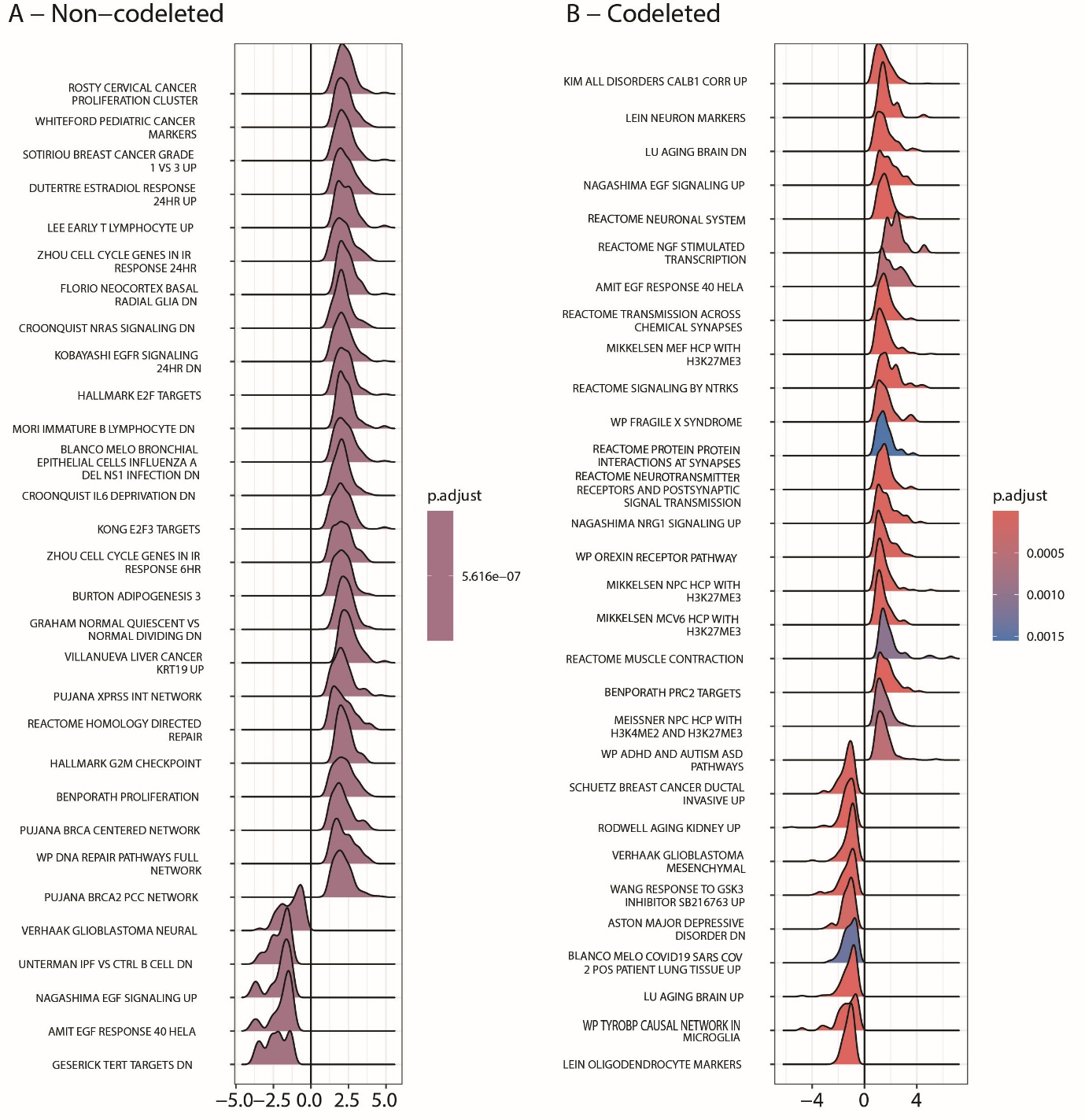

#### Figure S5. Top 30 pathways associated with TVGR in non-codel and codel LGG.

The top 30 pathways selected by GSEA are shown in a ridge-plots, illustrating the distributions of the z-scores.

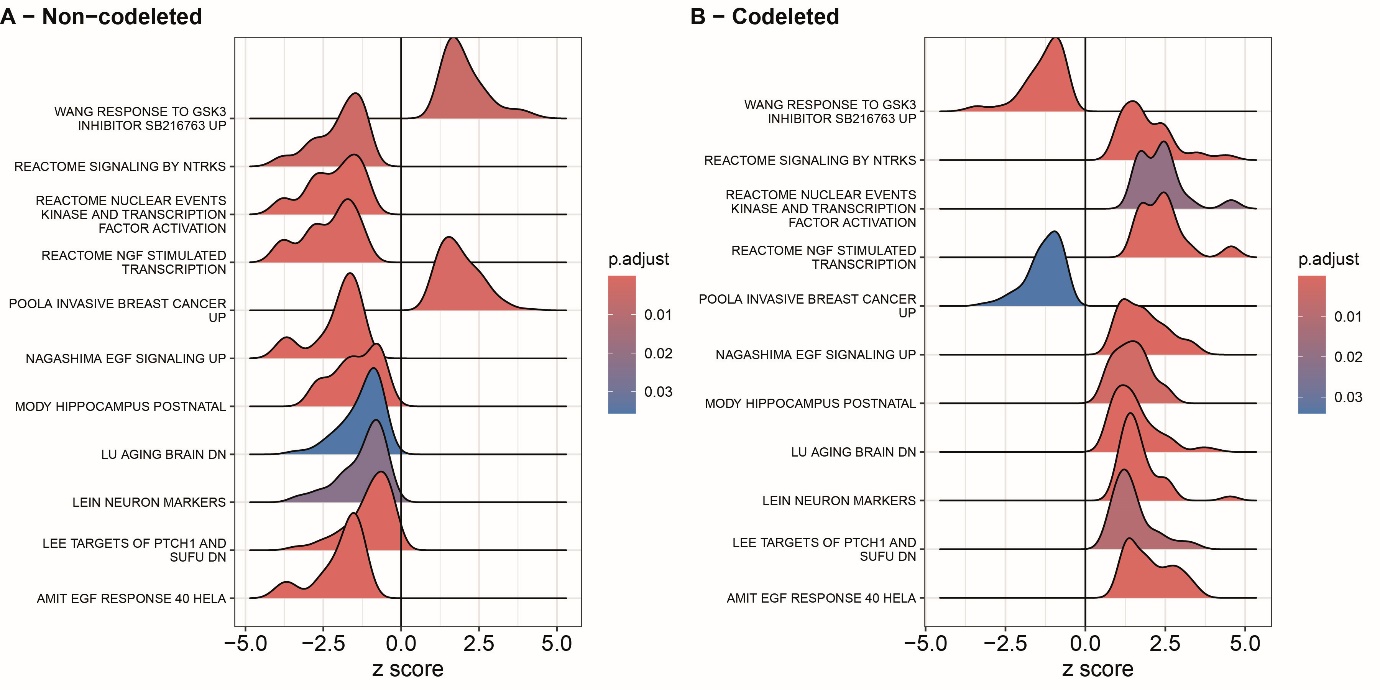

#### Figure S6. Common pathways associated with TVGR in non-codel and codel LGG.

The distributions of the z-scores are illustrated in the ridgeplot for the 11 common pathways selected by GSEA in non-codel (A) and codel (B) LGG. Of note, the significant association of the rank order has opposite signs.

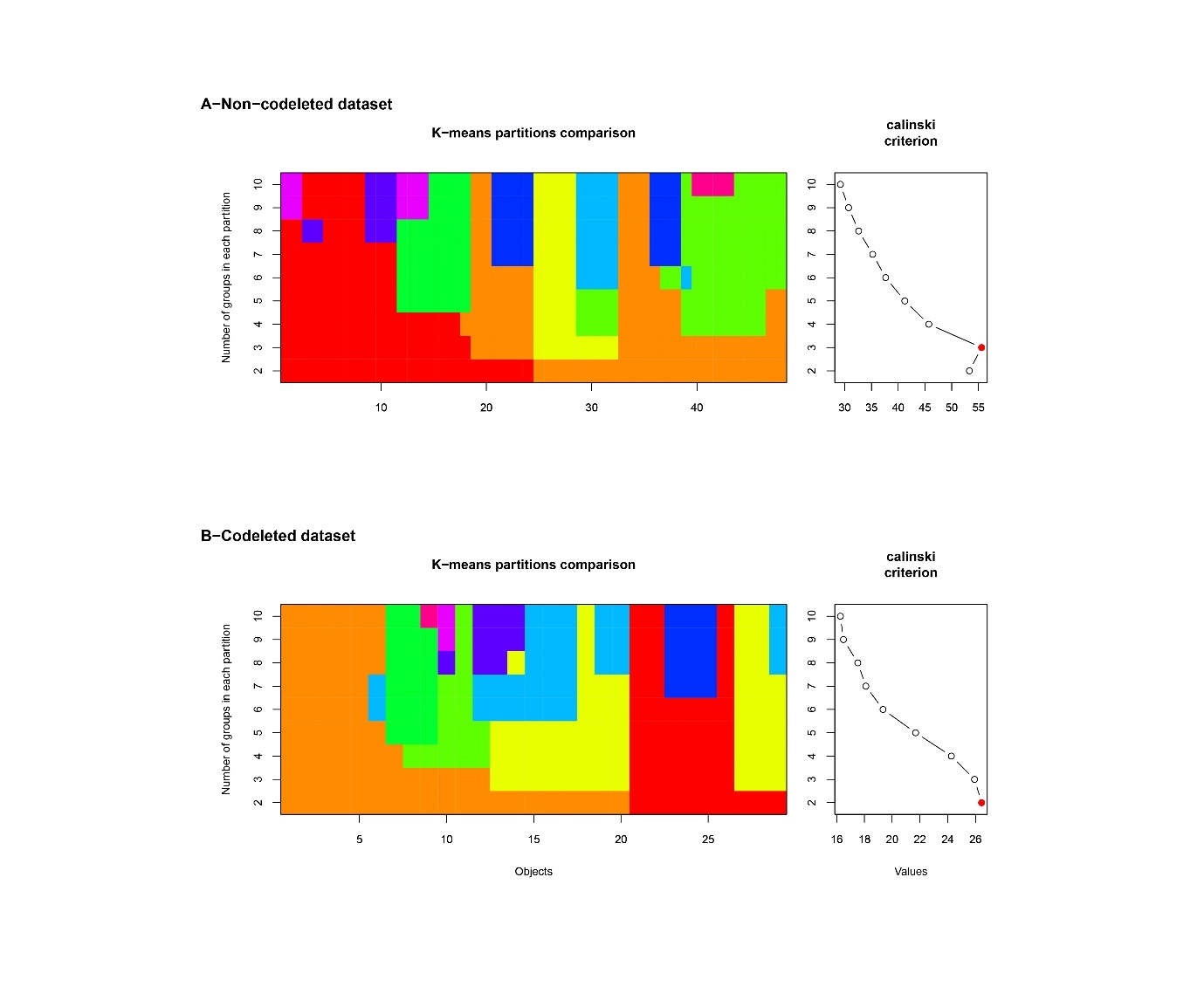

#### Figure S7. Cascade K Means partition comparison and Calinski plot versus number of clusters by LGG subtype.

Non-codel, **A**; codel, **B**. The color plots give the distribution of the samples in function of the groups defined by K-means. The calinski plot provides the optimal group number characterized by the higher values (red point). The optimal group number is 3 for IDHmt non-codel and 2 for codel LGG subtype.

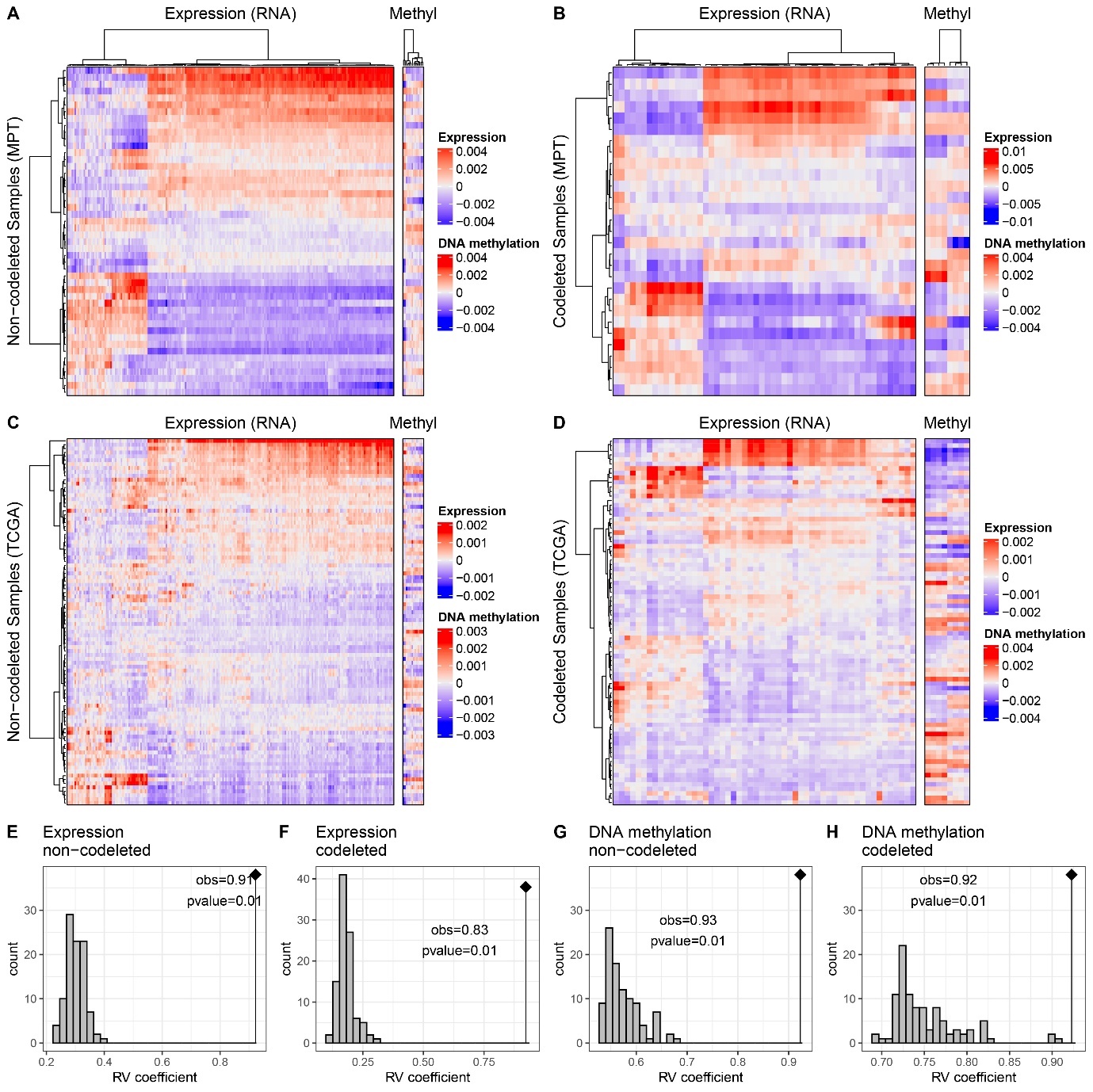

#### Figure S8. Comparison of TVGR associated pathway activities in the Montpellier cohorts to respective patterns in TCGA-LGG IDHmt grade II, stratified by LGG subtype (non-codel, codel).

The pathways are separated in two groups to show the contrast between pathways positively or negatively correlated with the TVGR as determined in the Montpellier cohorts. The heatmap for DNA methylation is based on the averaged gene expression and functional DNA methylation of the significant pathways emerging from GSEA (p ≤ 0.05). The DNA methylation and gene expression normalized datasets were weighted by their total inertia for the simultaneous heatmap representation as used in multiple factor analysis (MFA) beforehand. The dendrograms are based on Euclidean distance and Ward’s classification. The expression and DNA methylation structures are compared by RV coefficient tests based on 99 permutations for non-codeleted and codeleted datasets (E: expression for non-codeleted, F: expression for codeleted, G: DNA methylation for non-codeleted, H: DNA methylation for codeleted).

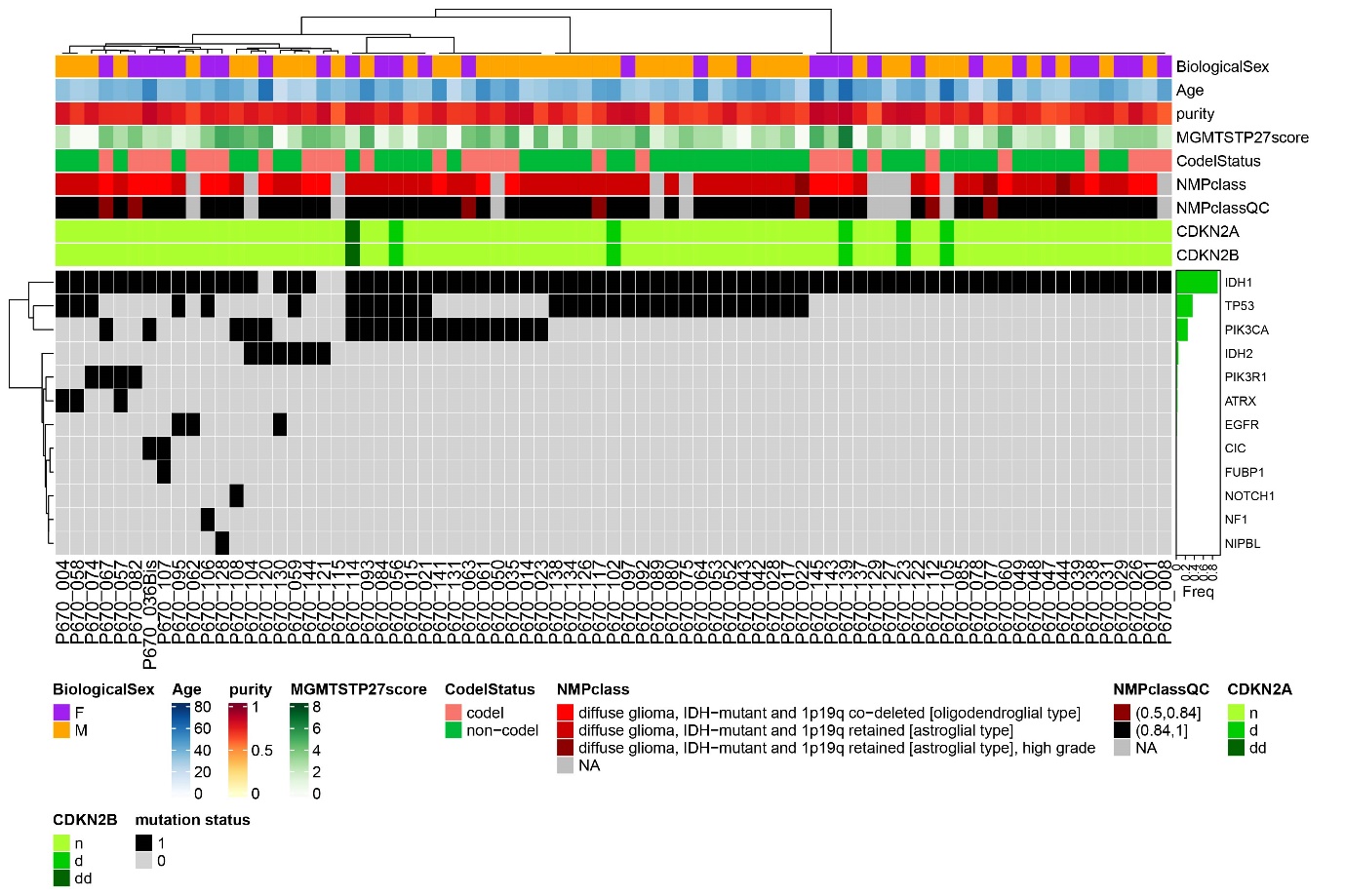

#### Figure S9. Mutation analysis for the non-codeleted and codeleted samples (n=77).

The RNA-seq data was used to determine mutations in the most commonly mutated genes reported from LGG and GBM in the COSMIC database (Tate et al., 2019). Additional sample annotations comprise: Biological sex, Age, Purity, MGMT-STP27 score, Codeletion status, NMPclass, NMPclassQC (Capper et al., 2018a; Capper et al., 2018b) and CNV of *CDKN2A/B*; n, normal; d, hemizygous deletion; dd, homozygous deletion.
